# Cognitive reserve as a moderator of the association between brain structure and later-life behavioural symptoms across the neurocognitive spectrum

**DOI:** 10.1101/2025.09.03.25335032

**Authors:** Gurshaan Sidhu, Dylan X. Guan, Maryam Ghahremani, Eric E. Smith, Zahinoor Ismail

## Abstract

**INTRODUCTION:** Cognitive reserve (CR) may buffer against the clinical manifestations of neurodegenerative disease. However, less is known about the relationship between CR and neuropsychiatric symptoms (NPS). Here, we investigated the moderation effect of CR on the relationship between AD-related cortical changes and NPS in older adults.

**METHODS:** Data were from 455 participants across the neurocognitive spectrum in the Comprehensive Assessment of Neurodegeneration and Dementia (COMPASS-ND) study. A composite CR score (CRS) was operationalized from measures of education, occupation, and mentally stimulating personal activities. The presence of clinically significant NPS was assessed using the Mild Behavioral Impairment Checklist. Logistic regression modelled the relationship between lower hippocampal volume or entorhinal thickness and NPS. A volume/thickness*CRS interaction term was included to assess effect modification. All models adjusted for age, sex, and cognitive status.

**RESULTS:** The association between lower hippocampal volume and higher odds of NPS was only present among participants with low CRS (OR: 1.46, 95%CI: [0.87, 2.46], p=.01), but not in participants with greater CRS (OR: 0.61, 95%CI: [0.39, 0.97], p=.01). A similar but weaker and non-significant interaction effect was observed with lower entorhinal thickness (high CRS, OR: 1.04, 95% CI: [0.52, 2.05], p=.34; low CRS, OR=1.59, 95% CI: [0.84, 2.97], p=.34).

**DISCUSSION:** Findings indicate that greater CR may mitigate NPS associated with lower hippocampal volume, independent of cognition, suggesting that CR may provide protective benefits beyond cognition. Future research should explore outcomes longitudinally, consider domain-specific analyses, and explore whether these findings extend to functional imaging surrogates of CR.

## INTRODUCTION

The concept of reserve explains why individuals with similar levels of brain health may differ in the extent of cognitive and functional impairment. The cognitive reserve (CR) theory proposes that individuals with efficient and well-integrated cognitive networks can better compensate for cognitive changes associated with brain damage, pathology, and aging (Stern, 2012). In other words, those with greater CR maintain better cognition despite having similar levels of brain pathology compared to individuals with lower CR. CR is thought to develop through lifelong experiences such as education, occupational complexity, and engagement in mentally stimulating activities (Stern et al., 2020). Studies have consistently shown that greater CR is associated with delayed onset of cognitive decline, particularly in the context of Alzheimer disease (AD) (Corbo et al., 2023). However, less is known about whether CR also buffers against non-cognitive outcomes, in the same way it does against cognition.

Neuropsychiatric symptoms (NPS), including apathy, affective dysregulation, impulse dyscontrol, social inappropriateness and psychosis are prevalent alongside cognitive symptoms in neurodegenerative diseases like AD. These symptoms are not only linked to poorer patient quality of life and function but also impose significant burden on caregivers (Fischer et al., 2012; Sheikh et al., 2018; Warring et al., 2024). Mild behavioral impairment (MBI), defined by later-life emergent and persistent NPS, identifies a high-risk group for incident dementia (Creese and Ismail, 2022; Ismail et al., 2016). MBI can manifest in individuals with normal cognition (NC), subjective cognitive decline (SCD), or mild cognitive impairment (MCI); at each of these cognitive stages, MBI is associated with a greater risk of incident cognitive decline and dementia (Ghahremani et al., 2025; Ismail et al., 2023; Ismail et al., 2021). A study of MCI participants showed that MBI predicted lower odds of reversion to NC, whereas transient NPS not meeting MBI criteria (non-MBI NPS) did not (McGirr et al., 2022). Therefore, MBI captures clinically meaningful NPS that represent the underlying neurodegenerative process.

Previous research has linked higher CR with lower odds of MBI using direct models of association (Guan et al., 2024a). However, the Collaboratory on Research Definitions for Reserve and Resilience in Cognitive Aging and Dementia has proposed that the ideal framework should demonstrate CR as a moderator of the relationship between brain status and clinical outcome (Stern et al., 2023). Atrophy or thinning of the brain regions associated with early AD pathology, such as the hippocampal and entorhinal cortices (Braak et al., 2006), is associated with MBI (Guan et al., 2024b; Matuskova et al., 2021). However, the moderating effect of CR on this relationship remains unknown. We investigated whether CR moderates the association between hippocampal and entorhinal cortices and the presence of clinically significant NPS in older adults across the neurocognitive spectrum. Based on CR theory, we hypothesized that individuals with greater CR will have a weaker association between lower volume or thickness in AD-related regions and the presence of NPS. Secondary objectives examined interactions in individuals without dementia, and of cognitive reserve subdomains.

## METHODS

### Study Design

The Comprehensive Assessment of Neurodegeneration and Dementia (COMPASS-ND) study is a longitudinal cohort study involving older adult participants with NC, SCD, MCI, or dementia (Chertkow et al., 2019). Participant data on clinical status, cognitive and behaviour assessments, demographics, lifestyle, and structural brain imaging are collected from multiple sites across Canada. A detailed description of the study has been published previously (Chertkow et al., 2019). Ethics approval was obtained from institutional ethics boards, and participant consent was obtained at each center affiliated with the study.

In this analysis, we used baseline cross-sectional data from 455 participants encompassing a neurocognitive spectrum from NC to MCI and dementia. The initial sample (n=632) comprised those who completed the Mild Behavioral Impairment Checklist (MBI-C), which was introduced partway through recruitment. Participants were excluded for age <50 years (n=2), missing data for the social network, support & activities questionnaire (n=18), hobbies and leisure activities questionnaire (n=3), employment history questionnaire (n=1), neurocognitive assessments (n=6), or structural magnetic resonance imaging (MRI) data (n=147) [Figure 1].

**Figure 1.**
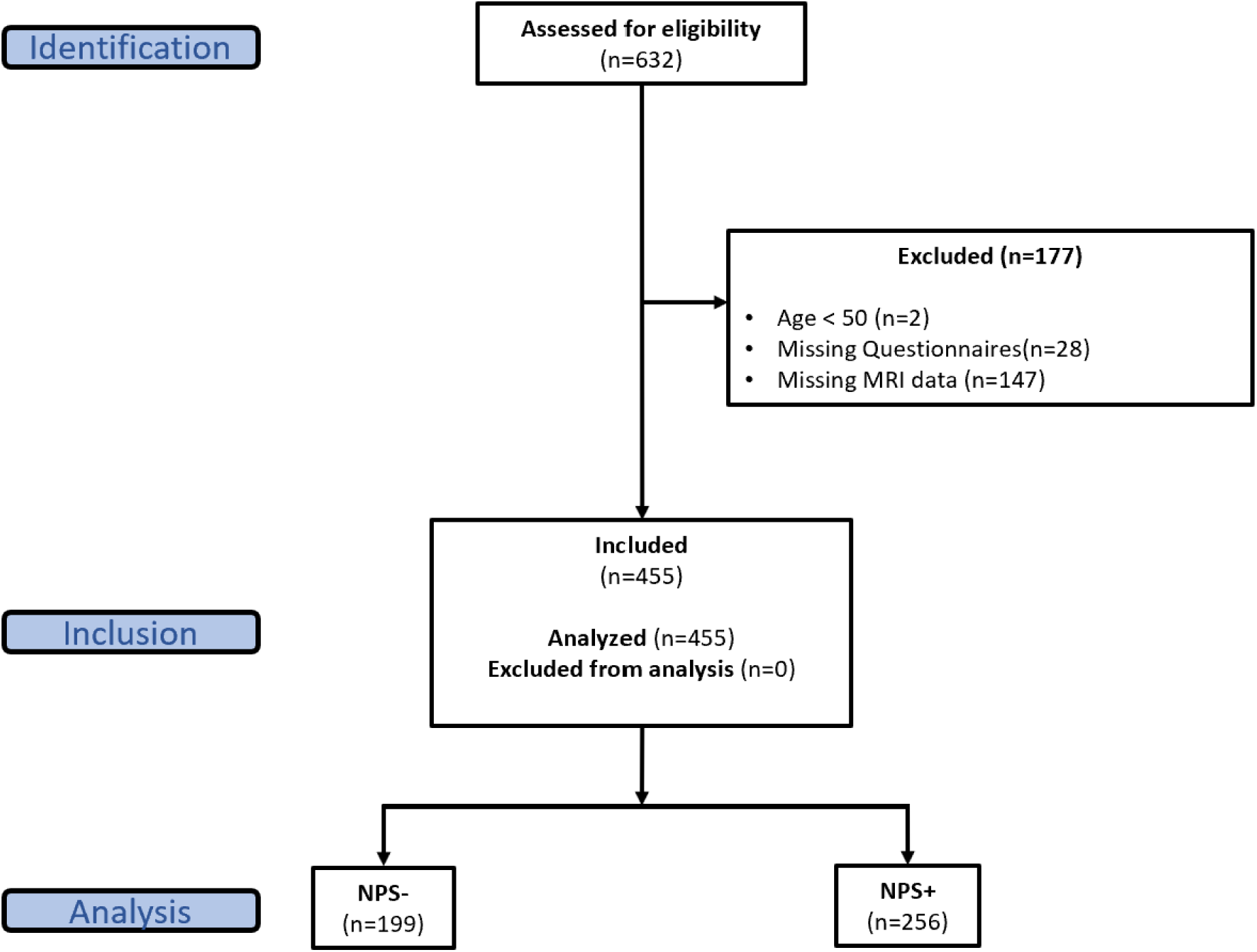
Participant Inclusion & Exclusion Flow Diagram. MBI - mild behavioral impairment, MBI-C – Mild Behavioural Impairment Checklist, MRI – magnetic resonance imaging.

### Cognitive Reserve Score Operationalization

A composite cognitive reserve score (CRS) was developed based on known proxy measures to quantify cognitive reserve across three domains: education (1 item), occupation (1 item), and personal activities (5 items) (Guan et al., 2024a; Stern, 2012) [Table 1]. Items were from self-reported and non-validated COMPASS-ND questionnaires. Educational attainment was measured by the highest level of education completed, as a more accurate proxy of cognitive reserve than total years of education (Harrison et al., 2015). Occupational complexity was assessed based on job type, with manual labor assigned the lowest complexity, and managerial roles assigned the highest complexity (Stern et al., 1994). The personal activities domain captured engagement in various cognitively stimulating activities, such as social interactions, multilingualism, and ability to play a musical instrument (Craik et al., 2010; Scarmeas et al., 2001). As shown in Table 1, items were coded such that a higher CRS indicated greater educational attainment, higher occupational complexity, or higher engagement in personal activities. CRS, measured using this approach, has been shown to be associated with neuropsychological test performance, lower odds of subjective decline, and lower self-reported cognitive symptoms in older adults (Guan et al., 2024a).

**Table 1.**
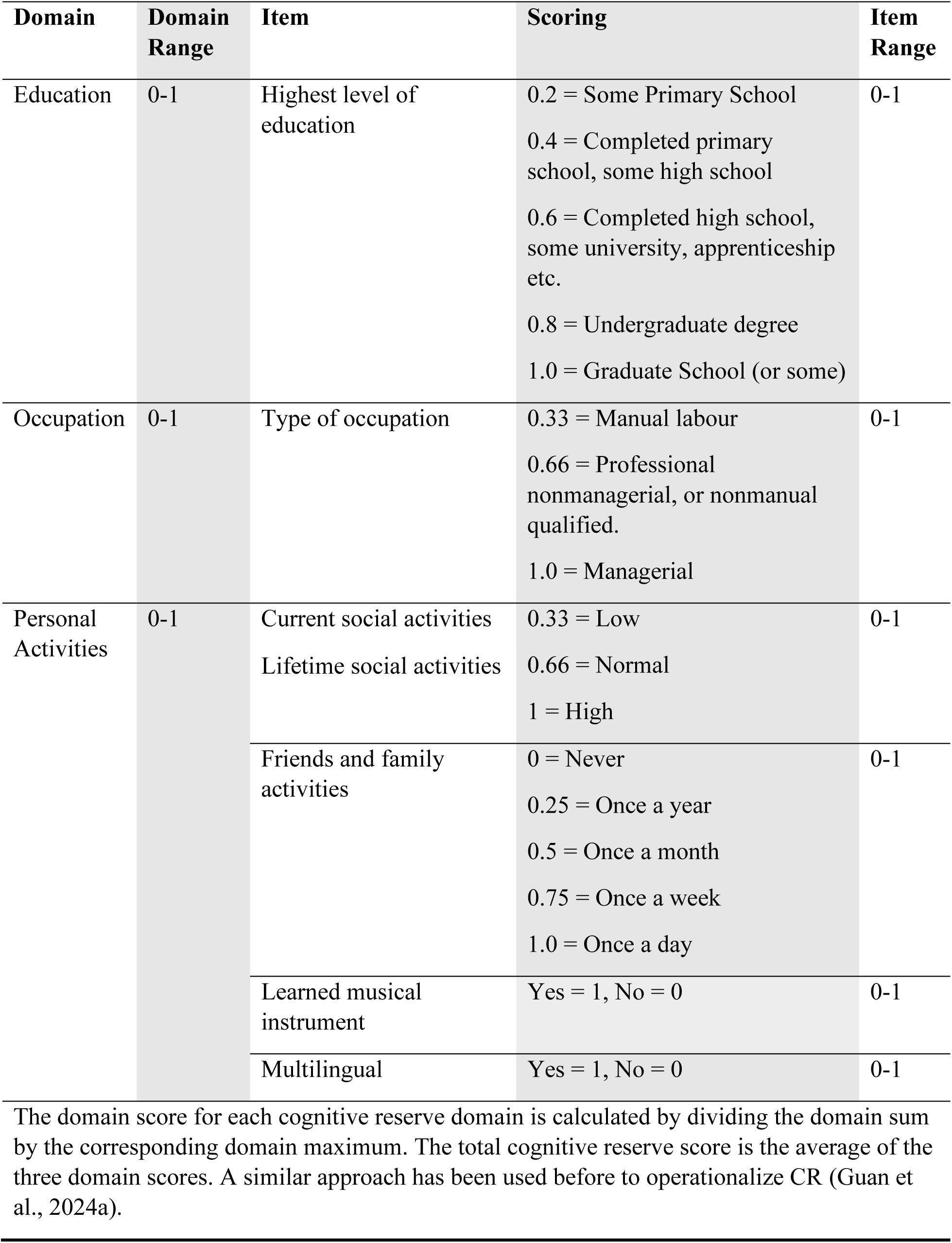
Cognitive Reserve Score Operationalization.

Each CRS domain had a range from 0-1, calculated by dividing the domain sum by the maximum total domain score. The total CRS was the mean of all three domain scores. We assumed equal weightage of the three domains, as their contribution to cognitive reserve are not yet established relative to each other.

### Cognitive Function

Neurocognitive status was categorized into three groups based on clinical diagnosis: (1) NC, including individuals with SCD, (2) MCI, including vascular and Parkinson’s disease-related MCI, and (3) dementia, including Alzheimer’s disease, Parkinson’s disease, frontotemporal, vascular, and Lewy body dementia. The Montreal Cognitive Assessment (MoCA) is a validated screening tool designed to assess various cognitive domains, including memory, attention, language, and executive function (Nasreddine et al., 2005). Higher CRS was expected to correlate with better cognitive function.

### NPS Assessment

The MBI-C is a rating scale designed to identify later-life emergent and persistent NPS in older adults in accordance with the MBI criteria (Ismail et al., 2017). The informant-rated MBI-C, which has been validated for use in dementia populations, was used for NPS assessment in our sample (Hu et al., 2023; Kassam et al., 2023). The MBI-C is comprised of 34-items split across five domains describing MBI symptoms: decreased motivation (6 items), affective dysregulation (6 items), impulse dyscontrol (12 items), social inappropriateness (5 items), and psychosis (5 items). Informants first confirmed whether the participant showed each symptom for at least six months, signifying a change from their usual behaviour. For symptoms that were present, the corresponding severity was rated using a scale from 1-3, with higher scores indicating greater symptom severity. Total MBI-C score was calculated by summing scores across all items (range = 0-102). Clinically significant NPS were classified using a total MBI-C cut-off score ≥ 6 (NPS+), consistent with previous studies in COMPASS-ND (Ghahremani et al., 2023).

### MRI acquisition and regions of interest

Structural MRI data, including 3D T1-weighted images, were collected for all participants and processed using standardized imaging protocols, generating thickness, surface area, and volume cortical maps (Chertkow et al., 2019; Duchesne et al., 2019). These measures were converted to Z scores adjusted for age, sex, estimated intracranial volume, scanner manufacturer, and magnetic field strength (Marcotte et al., 2019). Automated hippocampal and entorhinal segmentation was conducted by True Positive, a company specializing in real-time volumetrics for clinical trials (Chertkow et al., 2019).

Volumetric measures have been suggested to be inferior to thickness measures for measuring neurodegeneration in AD, particularly in samples with a broad age range (Schwarz et al., 2016). However, the hippocampal region is typically assessed through volumetric analysis due to limitations of thickness measurement techniques. Therefore, our analysis focused on hippocampal volume and entorhinal thickness as the regions of interest (ROIs), calculated as the average of standardized bilateral measures. ROI values were reverse coded so that higher scores corresponded to lower cortical volume or thickness.

### Statistical Analysis

R version 4.4.0 was used to conduct all statistical analyses. Participant characteristics, including demographics, clinical status, CRS, and cognitive and behaviour measures, were summarized for the entire sample and stratified by NPS group. Differences between the NPS+ and NPS-groups were assessed by independent samples t-tests for continuous variables or chi-square tests for discrete variables. Demographics of excluded and included participants were compared to identify possible demographic and clinical sources of self-selection bias.

Prior to testing our hypothesis, we first examined the convergent validity of CRS by modelling association with neurocognitive status (NC/SCD, MCI, dementia) and MoCA score, via ordinal logistic regression and linear regression models, respectively. For these models, we verified the relevant regression assumptions: for ordinal logistic regressions, we tested the proportional odds assumption using the Brant test; for linear regressions, we assessed normality of residuals and homoscedasticity through diagnostic plots. In addition to correlating with cognitive outcome, a valid proxy of CR should also moderate the association between brain pathology and cognitive performance (Stern et al., 2020). To test this, we assessed the moderation effect of CRS with cognitive status as the outcome by using ordinal logistic regression models with an interaction term between the ROI and CRS.

To test our hypothesis, a binomial logistic regression model was applied to the association between CRS and NPS status in our sample, thereby replicating prior studies on CRS and NPS (Guan et al., 2024a). Next, in alignment with the Collaboratory on Research Definitions for Reserve and Resilience in Cognitive Aging and Dementia (Stern et al., 2023), the moderation effect of CRS on the association between the ROIs and behavioural impairment was assessed via a binomial logistic regression model. For each ROI, the model included an interaction term between the ROI and CRS. A Johnson-Neyman/floodlight analysis was conducted to determine the range of CRS values at which the association between the ROI and NPS was statistically significant. Furthermore, considering that MBI represents an early clinical stage along the behavioural continuum, where the effects of CR are most evident (Stern, 2012), we analyzed it as a separate outcome than NPS status. To investigate the relationship between CRS and MBI, we analyzed a subsample of 355 participants without a diagnosis of dementia, as per MBI criteria (Ismail et al., 2016).

Secondary analyses explored interactions between the ROIs and each CRS subdomain (education, occupation, and personal activities) using the same statistical models described previously. Sex differences were tested by including sex in the interaction terms in each model. All statistical models were adjusted for participant age and sex. Cognitive status was controlled in models of NPS, and the presence of NPS was controlled in models of cognition to ensure that observed associations with CRS were not confounded by the relationship between cognition and behaviour. Statistical significance was set at a threshold of p<0.05.

## RESULTS

### Sample Characteristics

A comparison of participant demographics between those excluded and included in the study are shown in Supplementary Table 1. Participants that were excluded were more likely to be assigned male sex at birth (p<.001) and had more education (p=.04) and higher proportion of those with NC (p=.02). Study participant demographics and characteristics are shown in Table 2. Across all participants, 36.9% and 22.2% had MCI or dementia, respectively, and 43.7% were classified as NPS+. The study cohort was highly educated (15.2±2.7 years of education), with the NPS-group having more education than the NPS+ group (p=0.03). Compared to the NPS-group, the NPS+ group were more likely to be assigned male sex at birth (p<.001), had higher prevalence of MCI or dementia (p=.01), lower MoCA score (p<.001), and lower CRS (p=.01).

**Table 2.** Study Participant Characteristics.

| **Table 2. Study Participant Characteristics** |  |  |  |  |
| --- | --- | --- | --- | --- |
| **Variable** | **Total** | **NPS-** | **NPS+** | **p** |
| n | 455 | 256 | 199 |  |
| Age (years) | 70.6 (7.5), 50–91 | 70.0 (6.7), 53–91 | 71.3 (8.4), 50–89 | .06 |
| Sex (female) | 231 (50.7) | 148 (57.8) | 83 (41.7) | <.001 |
| Education (years) | 15.2 (2.7), 7–23 | 15.4 (2.5), 7–20 | 14.8 (2.9), 7–23 | .03 |
| Language (multilingual) | 274 (60.2) | 164 (64.1) | 110 (55.3) | .07 |
| MoCA Score | 23.3 (4.9), 4–30 | 25.2 (4.0), 13–30 | 21.5 (5.2), 4–30 | <.001 |
| Cognitive Status |  |  |  | <.001 |
| NC | 186 (40.9) | 145 (56.6) | 41 (20.6) |  |
| MCI | 168 (36.9) | 86 (33.6) | 82 (41.2) |  |
| Dementia | 101 (22.2) | 25 (9.8) | 76 (38.2) |  |
| Cognitive Reserve Score |  |  |  |  |
| Total | 0.68 (0.13), 0.25–1.00 | 0.70 (0.12), 0.31–1.00 | 0.65 (0.13), 0.25–0.95 | .01 |
| Education | 0.73 (0.18), 0.20–1.00 | 0.75 (0.18), 0.20–1.00 | 0.71 (0.18), 0.20–1.00 | .01 |
| Occupation | 0.68 (0.19), 0.33–1.00 | 0.70 (0.18), 0.33–1.00 | 0.65 (0.20), 0.33–1.00 | .007 |
| Personal | 0.63 (0.16), 0.17–1.00 | 0.65 (0.15), 0.23–1.00 | 0.59 (0.17), 0.17–1.00 | <.001 |
| MBI-C Severity Score |  |  |  |  |
| Global | 8.4 (11.4), 0–77.0 | 1.2 (1.6), 0–5.0 | 17.6 (11.9), 6.0–77.0 | <.001 |
| Decreased Motivation | 2.4 (3.4), 0–16.0 | 0.3 (0.8), 0–5.0 | 5.0 (3.7), 0–16.0 | <.001 |
| Affective Dysregulation | 2.3 (3.2), 0–17.0 | 0.4 (0.8), 0–4.0 | 4.7 (3.5), 0–17.0 | <.001 |
| Impulse Dyscontrol | 2.6 (4.3), 0–30 | 0.4 (0.8), 0–4.0 | 5.4 (5.2), 0–30 | <.001 |
| Social Inappropriateness | 0.6 (1.8), 0–15.0 | 0 (0.2), 0–1.0 | 1.4 (2.5), 0–15.0 | <.001 |
| Psychosis | 0.5 (1.5), 0–14.0 | 0 (0.2), 0–1.0 | 1.1 (2.1), 0–14.0 | <.001 |

Numeric variables are shown in mean (standard deviation), range. Categorical variables are shown in n (%). All values are rounded to one decimal place, except for p-values which are rounded to two or three decimals and the cognitive reserve score which is rounded to two decimals. NPS+ status is classified as MBI-C ≥ 6. Abbreviations: MBI-C, Mild Behavioral Impairment Checklist; MCI, mild cognitive impairment; MoCA, Montreal Cognitive Assessment; NC, normal cognition; NPS, neuropsychiatric symptoms.

### Cognitive Reserve and Cognition

The CRS showed convergent validity with both MoCA score and neurocognitive status. A 0.1 increase in CRS corresponded to a 0.8 point higher MoCA total score (95%CI: [0.48, 1.12], p<.001) and 17% lower odds of MCI or dementia (OR: 0.83, 95%CI: [0.72, 0.94], p=.005) [Table 3]. In ordinal regression models with cognitive status as the outcome, no significant interaction effects were observed between CRS and hippocampal volume or entorhinal thickness [Table 4]. Neither hippocampal volume (p=.61) nor entorhinal thickness (p=.76) interactions were modified by sex.

**Table 3.** CRS Associations with Cognition and NPS.

| **Table 3. CRS Associations with Cognition and NPS** |  |  |  |
| --- | --- | --- | --- |
| **Outcome Variable** | **B** | **95% CI** | **p** |
| MoCA Score | 0.80 | 0.48, 1.12 | <.001 |
| MoCA Score (subsample) | 0.71 | 0.44, 0.98 | <.001 |
|  | **OR** | **95% CI** | **p** |
| Cognitive Status | 0.83 | 0.72, 0.94 | .005 |
| Cognitive Status (subsample) | 0.75 | 0.62, 0.91 | .004 |
| NPS | 0.80 | 0.68, 0.94 | .007 |
| NPS (subsample) | 0.82 | 0.68, 0.98 | .03 |

Unstandardized coefficients (B) were estimated from linear regression models for MoCA score and represent change in MoCA score for a 0.1 increase in CRS. ORs were estimated from logistic binomial regression for NPS or ordinal logistic regression models for cognitive status and represent change in odds per 0.1 increase in CRS. The subsample refers to participants without dementia. All models controlled for age and sex, as well as cognitive or NPS status when appropriate. Abbreviations: CI, confidence interval; CRS, cognitive reserve score; MoCA, Montreal Cognitive Assessment; NPS, neuropsychiatric symptoms; OR, odds ratio.

**Table 4.**
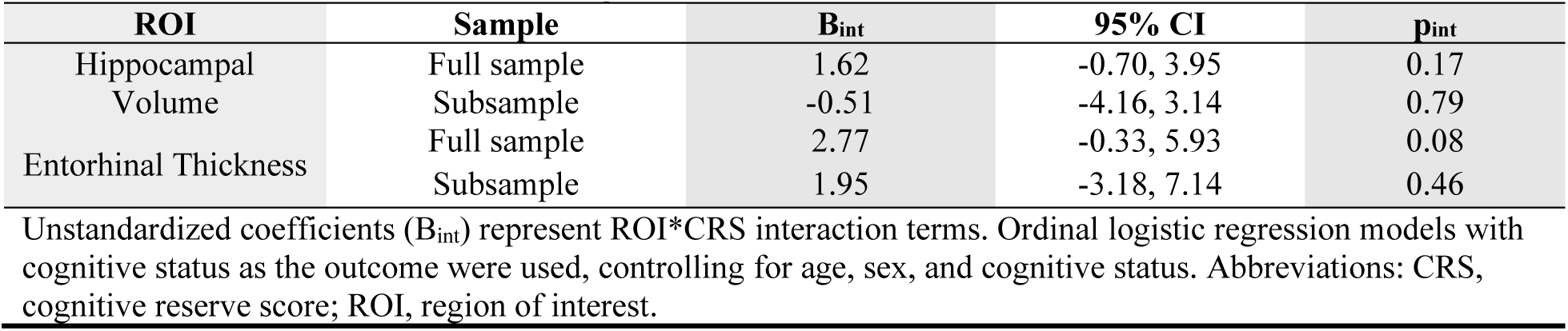
Interactions Terms for Cognitive Status Models.

| **ROI** | **Sample** | **B<sub>int</sub>** | **95% CI** | **p<sub>int</sub>** |
| --- | --- | --- | --- | --- |
| Hippocampal Volume | Full sample | 1.62 | -0.70, 3.95 | 0.17 |
|  | Subsample | -0.51 | -4.16, 3.14 | 0.79 |
| Entorhinal Thickness | Full sample | 2.77 | -0.33, 5.93 | 0.08 |
|  | Subsample | 1.95 | -3.18, 7.14 | 0.46 |

Unstandardized coefficients (B<sub>int</sub>) represent ROI\*CRS interaction terms. Ordinal logistic regression models with cognitive status as the outcome were used, controlling for age, sex, and cognitive status. Abbreviations: CRS, cognitive reserve score; ROI, region of interest.

### Cognitive Reserve and NPS

A 0.1 increase in CRS was associated with 18% lower odds of NPS+ (OR: 0.82, 95%CI: [0.68, 0.94], p=.007) status [Table 3]. This relationship was not modified by sex (p=.86). In logistic models with NPS as the outcome, the interaction term between CRS and hippocampal volume indicated that the association between hippocampal volume and NPS was significantly weaker at higher CRS score (B=-3.39, 95%CI: [−6.04, −0.84], p=.01) [Figure 2]. The models with an interaction term between the CRS and entorhinal thickness indicated a similar direction and magnitude of effect, but the estimate was less precise and non-significant (B=-1.65, 95%CI: [−5.10, 1.71], p=.34) [Figure 3]. Neither hippocampal volume (p=.73) nor entorhinal thickness (p=.17) interactions were modified by sex. Floodlight analysis revealed that the association between lower hippocampal volume and higher odds of NPS+ status was statistically significant (p<.05) when CRS was below 0.43.

**Figure 2.**
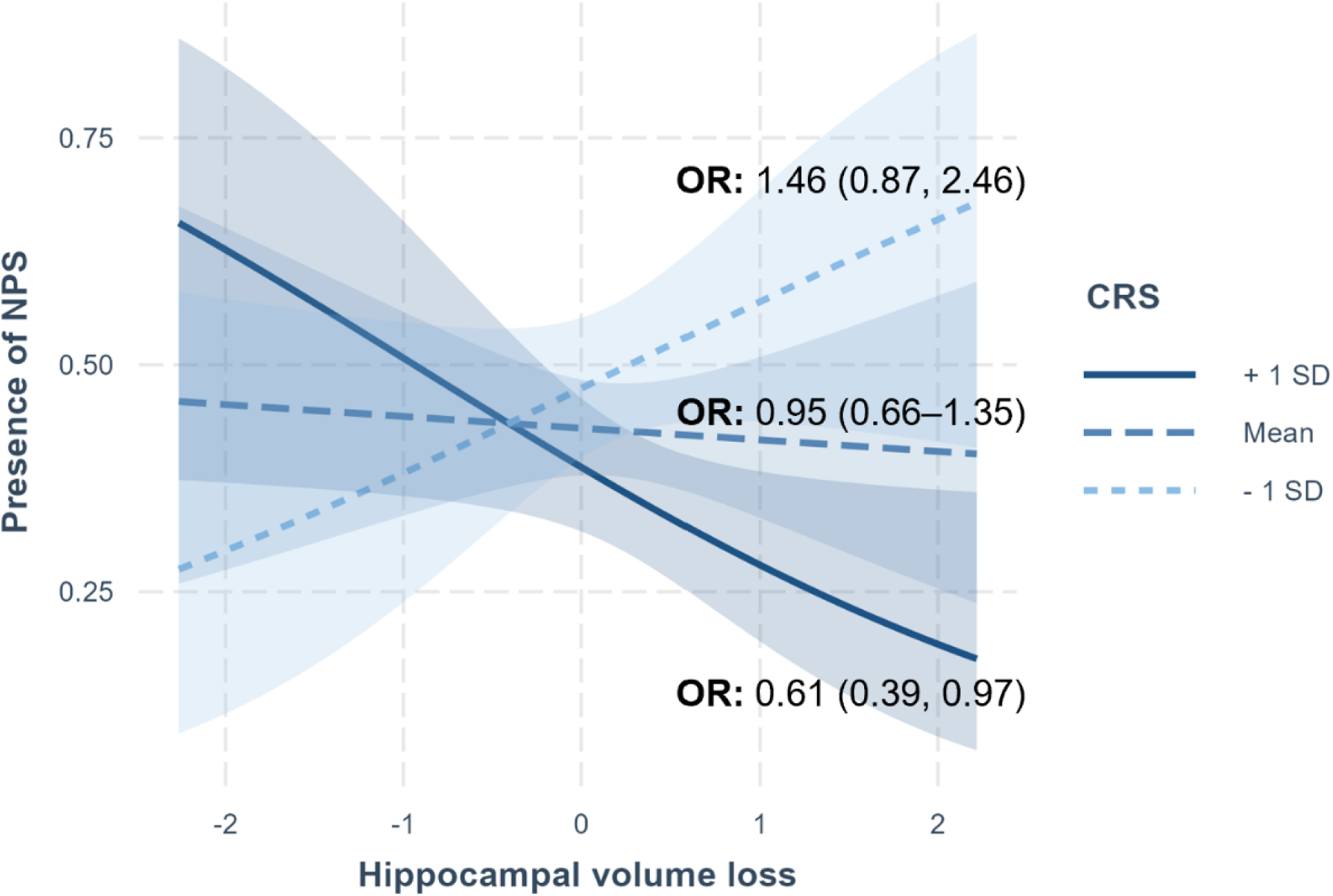
Marginal slopes depicting the association between hippocampal volume and NPS across different levels of CRS. ORs for NPS status are presented at low (−1 SD), average (mean), and high (+1 SD) CRS levels along with 95% CI in brackets. Logistic regression models with NPS as the outcome were used, controlling for age, sex, and cognitive status. Abbreviations: CI, confidence interval; CRS, cognitive reserve score; NPS, neuropsychiatric symptoms; OR, odds ratio.

**Figure 3.**
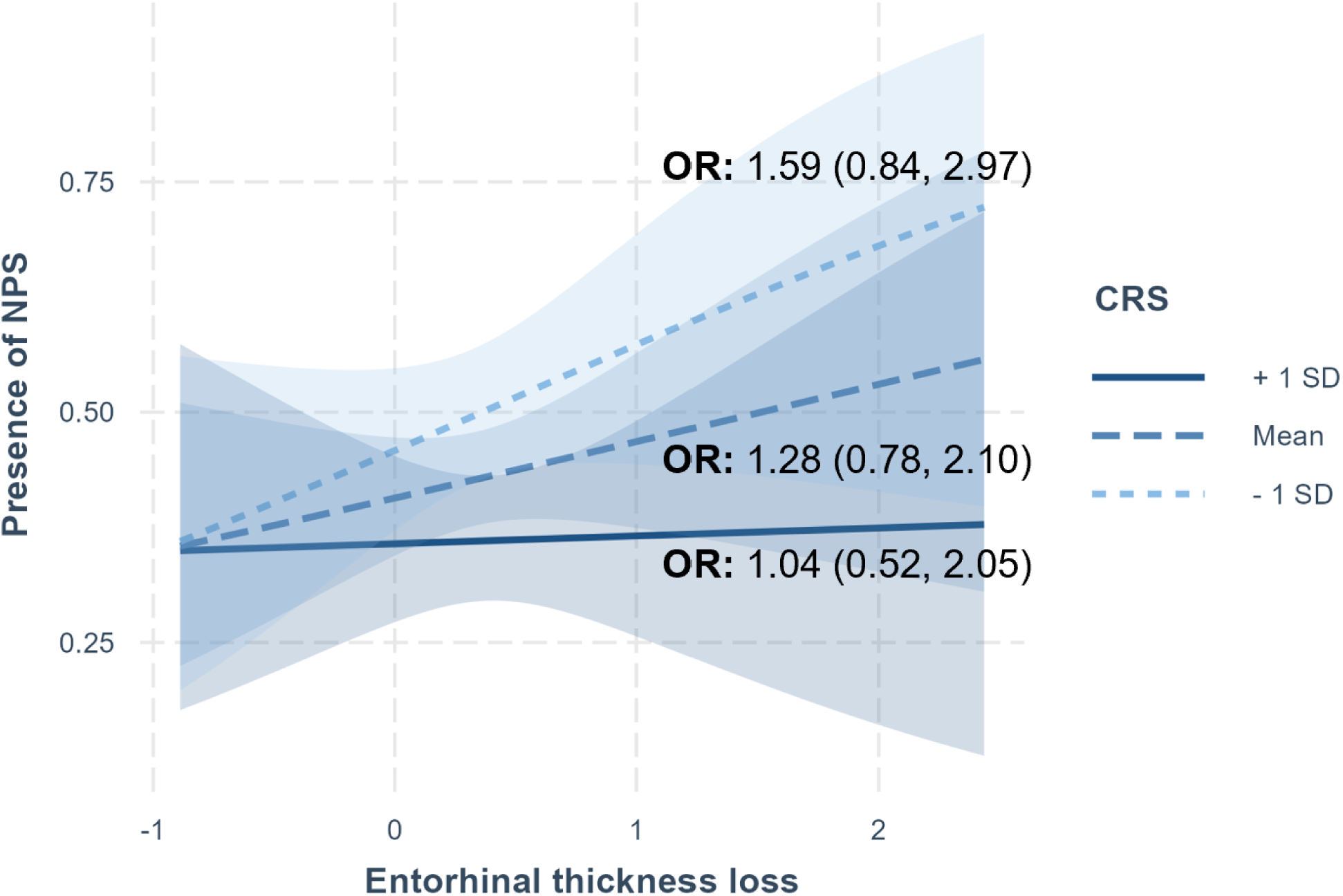
Marginal slopes depicting the association between entorhinal thickness and NPS across different levels of CRS. ORs for NPS status are presented at low (−1 SD), average (mean), and high (+1 SD) CRS levels along with 95% CI in brackets. Logistic regression models with NPS as the outcome were used, controlling for age, sex, and cognitive status. Abbreviations: CI, confidence interval; CRS, cognitive reserve score; NPS, neuropsychiatric symptoms; OR, odds ratio.

### Secondary Analyses

Participant demographics and characteristics of those without dementia are shown in Supplementary Table 2. In this subsample, in models of cognition, interaction effects for CRS and both hippocampal volume and entorhinal thickness on cognitive status were not observed [Table 4]. In models of NPS, the interaction effect between CRS and hippocampal volume was larger (B=-4.32, 95%CI: [−7.84, −1.04], p=.01) than in the full sample [Supplementary Figure 1]. The interaction with entorhinal thickness had a stronger effect size in the same direction as the full sample, but remained non-significant (B=-3.57, 95%CI: [−8.51, 0.87], p=.13) [Supplementary Figure 2]. Floodlight analysis demonstrated that the association between lower hippocampal volume and higher odds of MBI status was statistically significant (p<.05) when CRS was below 0.53.

To explore the contributions of CRS domains, secondary analyses examined interactions between hippocampal volume and the three CRS subdomains: education, occupation, and personal activities. The interaction between hippocampal volume and education was significant and had the strongest effect (B=2.87, 95%CI: [1.00, 4.82], p=.003). The interaction with the occupation domain was non-significant and had a slightly smaller effect (B=1.65, 95%CI: [−0.07, 3.44], p=.06). The personal activities domain had a similar effect size, which was also non-significant (B=1.40, 95%CI: [−0.76, 3.59], p=.20) [Supplementary Table 3 for all CRS subdomain interactions].

## DISCUSSION

In a sample of 455 older adults across the neurocognitive spectrum, an interaction effect was observed between CR and cortical ROIs, in association with behavioural outcomes, aligning with the expected CR framework. Specifically, the association between lower hippocampal volume (suggestive of AD-related atrophy) and presence of NPS was only present in those with low CRS; the association weakened with higher CRS. Moreover, this effect was stronger in participants without dementia, suggesting a stronger role of behavioural reserve in earlier stages of neurodegenerative disease, during which NPS i.e., behavioural and psychological symptoms of dementia (BPSD) may be classified as MBI. This finding aligns with the idea that CR is most effective before compensatory mechanisms are overwhelmed (Stern, 2012). While a similar interaction effect was seen between lower entorhinal thickness and NPS, it did not reach statistical significance. Additionally, no significant moderation effect between was observed when cognitive status was the outcome.

The moderation effect of CR observed for NPS suggests that the protective effect of CR may extend to non-cognitive outcomes, which is informative for future clinical and research practices. Along with mitigating cognitive decline, CR may also protect against BPSD, MBI, and the associated adverse health outcomes such as greater caregiver burden, institutionalization, frailty, and lower quality of life (Afram et al., 2014; Guan et al., 2022; Sheikh et al., 2018; Warring et al., 2024). Therefore, clinicians may be more likely to emphasize activities that build CR, knowing that it links to both cognitive and behavioural symptoms. Furthermore, these findings suggest the existence of common neurobiological substrates underlying both cognition and behaviour, setting the stage for future research to identify these substrates as more effective therapeutic targets that address multiple symptoms.

While behavioural symptoms are increasingly recognized as early clinical manifestations of neurodegenerative disease, with MBI being an early manifestation of BPSD, the behavioural dimension of reserve is less understood. Previous research has focused on patients with AD, and found more education to be associated with less apathy, depression, and irritability (Apostolova et al., 2014; Lobo et al., 1995; Zhao et al., 2016). Subsequently, similar associations were found in pre-dementia populations, specifically in persons with amnestic MCI (Inamura et al., 2022). More recently, a study with frontotemporal dementia patients showed that those with higher educational attainment can tolerate greater brain pathology in regions associated with disinhibited behaviour (Premi et al., 2013). Furthermore, a composite sociobehavioural measure of CR (incorporating educational attainment, type of occupation, and personal activities) has been linked to lower odds of MBI (Guan et al., 2024a).

However, some older studies found no association, or even a positive relationship, between education and NPS, contrasting with the behavioural reserve concept (Binetti et al., 1993; Gilley et al., 1991). Notably, these studies were limited by a small sample size (n<100) and potential methodological factors. For instance, NPS have often been measured without differentiating between longstanding psychiatric conditions, transient symptoms related to life stressors, and emergent and persistent behavioural changes related to neurodegenerative disease processes. Indeed, these studies lacked a validated approach to measure NPS or measured only one NPS domain. The use of MBI-C to measure NPS then represents a strength of our study, as it is designed to capture later-life emergent and persistent symptoms, enhancing specificity for neurodegenerative etiology (Ismail et al., 2017). The greater reliability and specificity of the MBI-C likely enhanced the detection of associations with NPS in this study. Additionally, this study also differed in the operationalization of CR by incorporating multiple domains, and in utilizing a sample that was representative across the neurocognitive spectrum.

Our CRS, operationalized using measures of education, occupation, and personal activities, demonstrated convergent validity with cognitive performance and behavioural symptoms. Our results also suggested that education was the most significant CRS domain. These findings are consistent with prior studies, suggesting that early-life reserve factors may offer greater protection against later-life cognitive and behavioural changes (Guan et al., 2024a). Though, the relative contribution of each CR domain remains unclear and thus warrants further investigation.

Although CR could possibly influence the atrophy-NPS association by buffering against cognitive decline, this is likely not the primary mechanism involved. The moderation effect of CR in this study remained significant, despite controlling for neurocognitive status, indicating that this effect was independent of cognition. This is consistent with previous studies on behavioural reserve that also adjusted for cognitive impairment (Apostolova et al., 2014; Guan et al., 2024a; Premi et al., 2013).

Instead, the observed moderation effect may be better explained by underlying mechanisms such as neural reserve or compensation. Neural reserve refers to the utilization of efficient and flexible brain networks, while neural compensation involves the recruitment of alternative networks to perform a task (Steffener and Stern, 2012). The default mode network (DMN) is one such network, implicated in both cognitive and emotional processes (Anthony and Lin, 2017; Smallwood et al., 2021). MBI has been associated with disrupted functional connectivity within the DMN; this connectivity change is proposed as a potential mediating pathway between CR and MBI (Ghahremani et al., 2023; Guan et al., 2024a). The hippocampus is a central component of the DMN, and while the entorhinal cortex is also frequently associated with the DMN, it is less directly involved (Raichle, 2015).

The hippocampi and entorhinal cortices are amongst the earliest and most affected regions in AD. Unlike the hippocampus, however, the moderation effect of entorhinal cortex did not meet statistical significance. This is consistent with the idea that hippocampal volume, despite being affected later in AD progression, is a more reliable measure of AD pathology than entorhinal thickness (Schwarz et al., 2016). Another possible explanation for this discrepancy is the differing degree of integration each region has with the DMN, though this remains speculative and needs further investigation.

Future research should incorporate functional MRI to better elucidate the neural mechanisms underlying CR. Additionally, future studies should investigate associations between CR and MBI severity, including analyses of MBI subdomains. Finally, longitudinal designs are needed to explore how CR influences NPS over time.

## Data Availability

Interested researchers may contact to request access to the data.

## Supplementary Tables & Figures

**Supplementary Figure 1.**
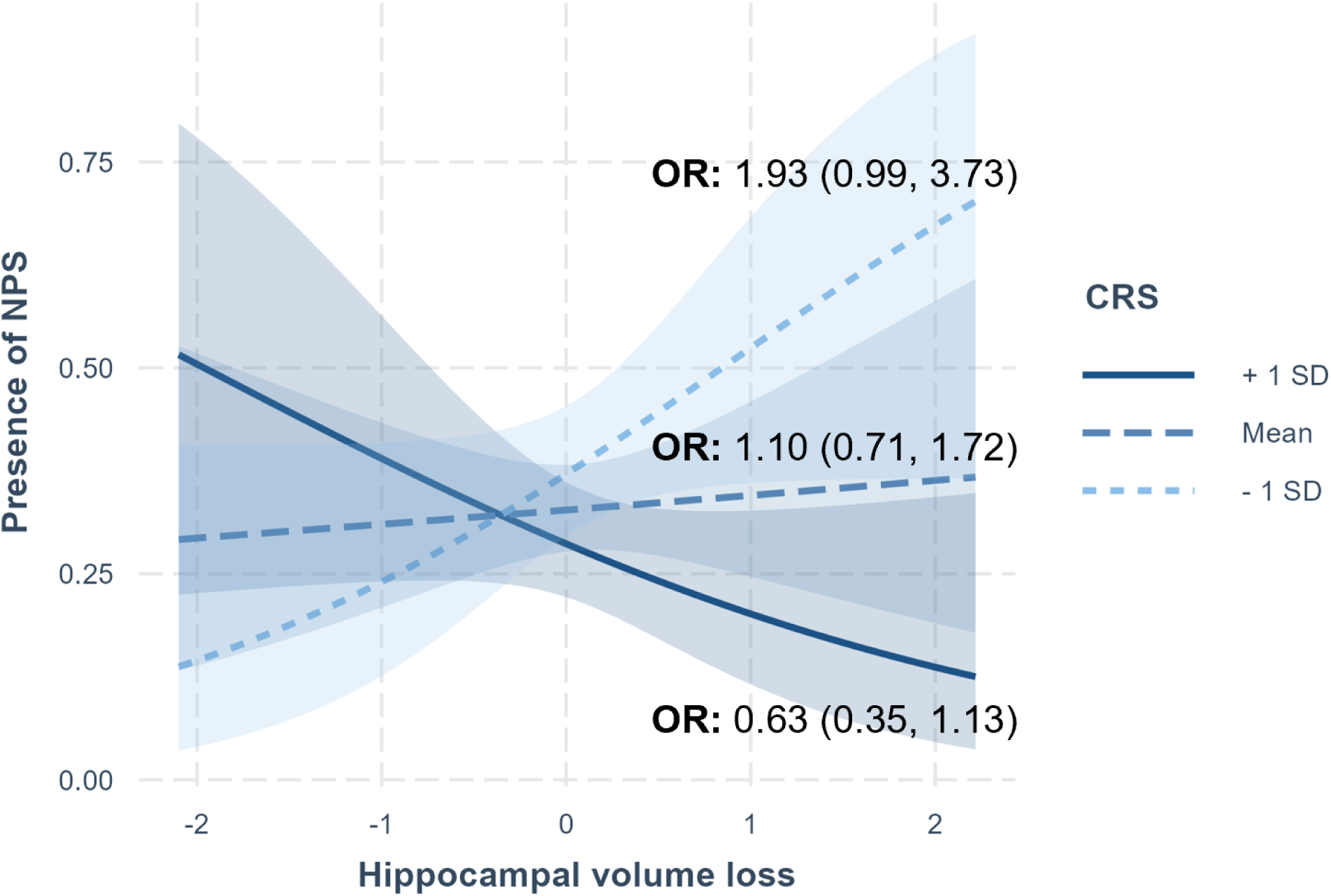
Marginal slopes depicting the association between hippocampal volume and NPS across different levels of CRS in those without dementia. ORs for NPS status are presented at low (−1 SD), average (mean), and high (+1 SD) CRS levels along with 95% CI in brackets. Logistic regression models with NPS as the outcome were used, controlling for age, sex, and cognitive status. Abbreviations: CI, confidence interval; CRS, cognitive reserve score; NPS, neuropsychiatric symptoms; OR, odds ratio.

**Supplementary Figure 2.**
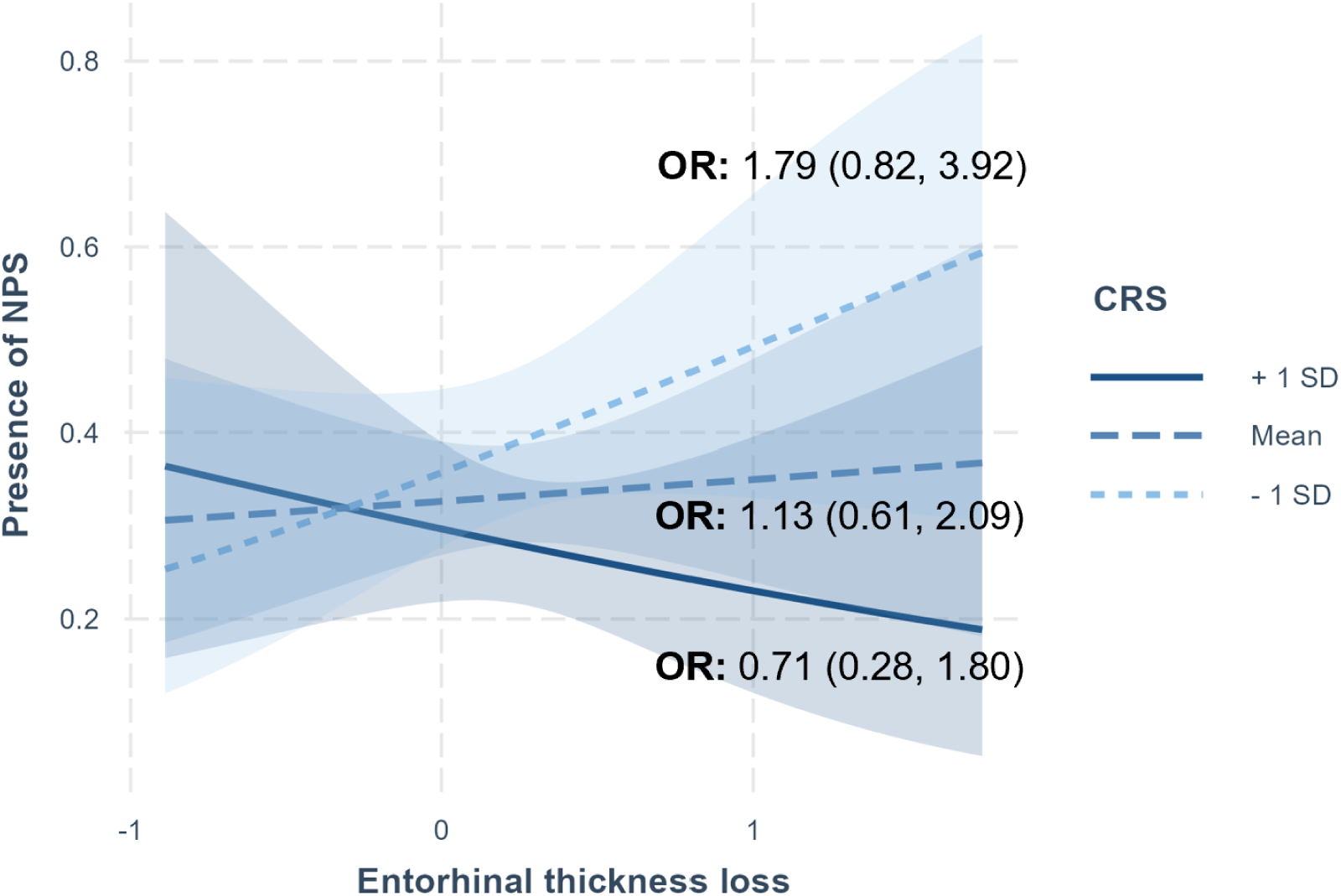
Marginal slopes depicting the association between entorhinal thickness and NPS across different levels of CRS in those without dementia. ORs for NPS status are presented at low (−1 SD), average (mean), and high (+1 SD) CRS levels along with 95% CI in brackets. Logistic regression models with NPS as the outcome were used, controlling for age, sex, and cognitive status. Abbreviations: CI, confidence interval; CRS, cognitive reserve score; NPS, neuropsychiatric symptoms; OR, odds ratio.

**Table S1.**
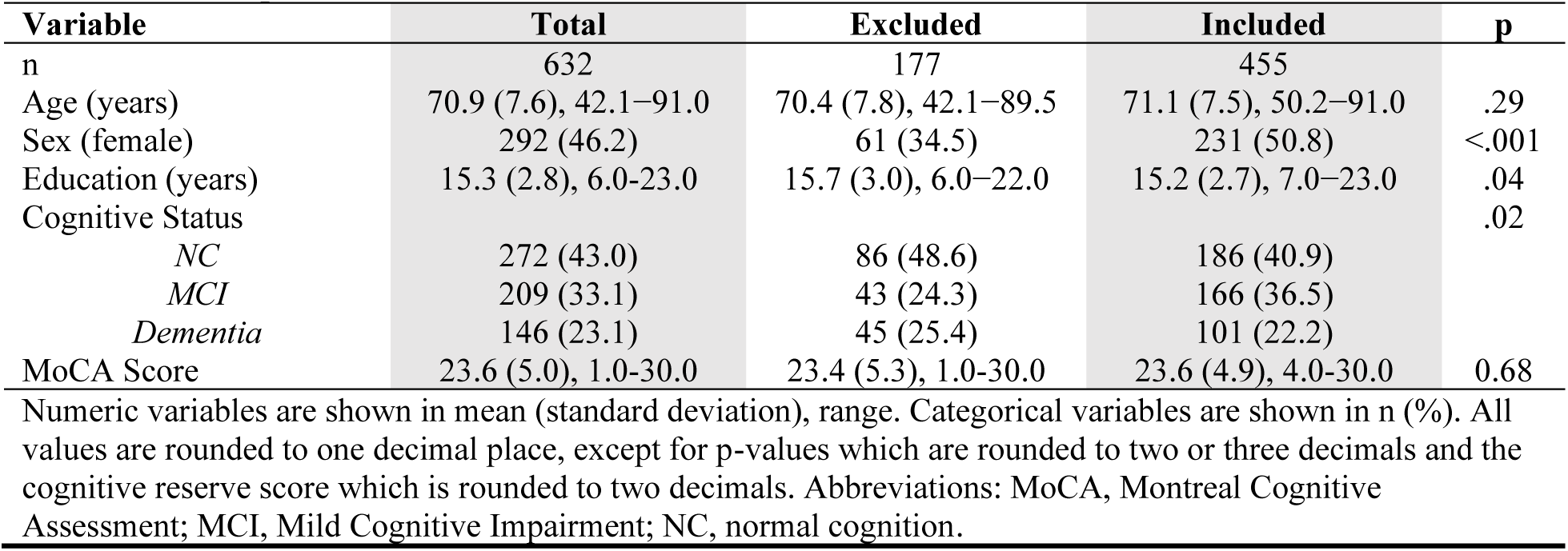
Participant Inclusion & Exclusion Table.

Supplementary Tables & Figures
| **Table S1. Participant Inclusion & Exclusion Table** |  |  |  |  |
| --- | --- | --- | --- | --- |
| **Variable** | **Total** | **Excluded** | **Included** | **p** |
| n | 632 | 177 | 455 |  |
| Age (years) | 70.9 (7.6), 42.1–91.0 | 70.4 (7.8), 42.1–89.5 | 71.1 (7.5), 50.2–91.0 | .29 |
| Sex (female) | 292 (46.2) | 61 (34.5) | 231 (50.8) | <.001 |
| Education (years) | 15.3 (2.8), 6.0–23.0 | 15.7 (3.0), 6.0–22.0 | 15.2 (2.7), 7.0–23.0 | .04 |
| Cognitive Status |  |  |  | .02 |
| NC | 272 (43.0) | 86 (48.6) | 186 (40.9) |  |
| MCI | 209 (33.1) | 43 (24.3) | 166 (36.5) |  |
| Dementia | 146 (23.1) | 45 (25.4) | 101 (22.2) |  |
| MoCA Score | 23.6 (5.0), 1.0–30.0 | 23.4 (5.3), 1.0–30.0 | 23.6 (4.9), 4.0–30.0 | 0.68 |

Numeric variables are shown in mean (standard deviation), range. Categorical variables are shown in n (%). All values are rounded to one decimal place, except for p-values which are rounded to two or three decimals and the cognitive reserve score which is rounded to two decimals. Abbreviations: MoCA, Montreal Cognitive Assessment; MCI, Mild Cognitive Impairment; NC, normal cognition.

**Table S2.** Non-Dementia Participant Characteristics.

| **Table S2. Non-Dementia Participant Characteristics** |  |  |  |  |
| --- | --- | --- | --- | --- |
| **Variable** | **Total** | **MBI-** | **MBI+** | **p** |
| n | 354 | 231 | 123 |  |
| Age (years) | 69.9 (7.2), 50–91 | 69.6 (6.6), 53–91 | 70.5 (8.2), 50–89 | .27 |
| Sex (female) | 191 (54.0) | 138 (59.7) | 53 (43.1) | .004 |
| Education (years) | 15.2 (2.7), 7–23 | 15.4 (2.5), 7–20 | 14.9 (3.0), 7.0–23 | .07 |
| Language (multilingual) | 218 (61.6) | 145 (62.8) | 73 (59.3) | .61 |
| MoCA Score | 25.3 (3.5), 13–30 | 26.0 (3.2), 18–30 | 24.1 (3.9), 13–30 | <.001 |
| Cognitive Status |  |  |  | <.001 |
| NC | 186 (52.5) | 145 (62.8) | 41 (33.3) |  |
| MCI | 168 (47.5) | 86 (37.2) | 82 (66.7) |  |
| Cognitive Reserve Score |  |  |  |  |
| Total | 0.67 (0.13), 0.28–0.96 | 0.69 (0.12), 0.28–0.96 | 0.65 (0.14), 0.31–0.96 | .003 |
| Education | 0.74 (0.18), 0.20–1.00 | 0.75 (0.18), 0.20–1.00 | 0.72 (0.19), 0.20–1.00 | .09 |
| Occupation | 0.68 (0.18), 0.33–1.00 | 0.70 (0.18), 0.33–1.00 | 0.65 (0.20), 0.33–1.00 | .02 |
| Personal | 0.59 (0.16), 0.13–1.00 | 0.61 (0.15), 0.18–0.95 | 0.57 (0.17), 0.13–1.00 | .02 |
| MBI-C Severity |  |  |  |  |
| Global | 6.6 (10.7), 0–77.0 | 1.0 (1.5), 0–5.0 | 17.1 (12.5), 6.0–77.0 | <.001 |
| Decreased Motivation | 1.7 (3.0), 0–16.0 | 0.2 (0.6), 0–4.0 | 4.4 (3.7), 0–16.0 | <.001 |
| Affective Dysregulation | 1.8 (3.0), 0–17.0 | 0.4 (0.8), 0–4.0 | 4.6 (3.6), 0–17.0 | <.001 |
| Impulse Dyscontrol | 2.2 (4.0), 0–30 | 0.4 (0.8), 0–4.0 | 5.6 (5.2), 0–30 | <.001 |
| Social Inappropriateness | 0.5 (1.6), 0–15.0 | 0 (0.2), 0–1.0 | 1.5 (2.5), 0–15.0 | <.001 |
| Psychosis | 0.4 (1.4), 0–14.0 | 0 (0.1), 0–1.0 | 1.1 (2.2), 0–14.0 | <.001 |

Numeric variables are shown in mean (standard deviation), range. Categorical variables are shown in n (%). All values are rounded to one decimal place, except for p-values which are rounded to two or three decimals and the cognitive reserve score which is rounded to two decimals. NPS+ status is classified as MBI-C ≥ 6. Abbreviations: MBI, mild behavioral impairment; MBI-C, Mild Behavioral Impairment Checklist; MCI, mild cognitive impairment; MoCA, Montreal Cognitive Assessment; NC, normal cognition.

**Table S3.** Interactions Terms between Hippocampal Volume and CRS Domains.

| CRS Domain | Sample | B<sub>int</sub> | 95% CI | p<sub>int</sub> |
| --- | --- | --- | --- | --- |
| Education | Full Sample | 2.87 | 1.00, 4.82 | **.003** |
|  | Subsample | 3.10 | 0.85, 5.52 | **.009** |
| Occupation | Full Sample | 1.65 | -0.07, 3.44 | .06 |
|  | Subsample | 2.12 | -0.18, 4.54 | .08 |
| Personal Activities | Full Sample | 1.40 | -0.76, 3.59 | 0.20 |
|  | Subsample | 2.26 | -0.65, 5.28 | 0.13 |

Unstandardized coefficients (B<sub>int</sub>) represent hippocampal volume\*CRS domain interaction terms. Logistic regression models with NPS as the outcome were used, controlling for age, sex, and cognitive status.
Abbreviations: CI, confidence interval; CRS, cognitive reserve score; NPS, neuropsychiatric symptoms.

